# A large-scale single cell map of primary and conditional regulatory variation in the human brain

**DOI:** 10.64898/2026.05.30.26354534

**Authors:** Sungbeen Lee, Zane Hamdan, Sommer S. Huntress, Amy Yang, Michael Guo

**Author notes:** equal contributions (Sungbeen Lee, Zane Hamdan).

## Abstract

Most disease-associated genetic variants reside in noncoding regions and are thought to influence risk by altering gene regulation. However, linking these variants to target genes and mechanisms remains challenging because regulatory effects can be cell type specific and may involve multiple independent signals at the same locus. Although eQTL studies have identified many regulatory variants, many GWAS loci do not colocalize with primary eQTLs, suggesting that additional regulatory signals remain unresolved. Here, we generated a large-scale single cell genomic resource to map primary and conditional regulatory variation in the human brain. We integrated single nucleus RNA-seq data from 956 postmortem brains, comprising 4.8 million nuclei across seven major brain cell types, with matched whole genome sequencing data. Using data from a subset of 763 individuals of European ancestry, we identified single cell eQTLs (sc-eQTLs) for 32,189 eGenes across cell types and uncovered 6,174 additional conditionally independent sc-eQTL signals through stepwise conditional analysis. Conditional sc-eQTLs were more cell type-specific than primary sc-eQTLs and showed distinct regulatory architectures, including greater distances from TSSs and reduced enrichment in annotated promoters and enhancers. Genes with conditional sc-eQTLs also showed lower genic constraint in most cell types, suggesting that genes with secondary regulatory effects are shaped by different selective pressures. Importantly, several schizophrenia and Alzheimer’s disease GWAS loci colocalized more strongly with conditional than primary sc-eQTLs, demonstrating that secondary regulatory signals can reveal disease-relevant mechanisms missed by primary sc-eQTL analyses alone. Together, these findings show that conditional and cell type-specific regulatory variation represents a substantial component of the genetic architecture of brain disease.

## Introduction

Most genome-wide association study (GWAS) associations map to noncoding regions of the genome and are thought to influence disease risk through regulation of gene expression^1^. Despite the success of GWAS in identifying thousands of disease-associated loci, linking noncoding variants to the precise regulatory mechanisms and the target genes they regulate remains a major challenge^2,3^. This challenge reflects several features of human genome organization and gene regulation. First, extensive linkage disequilibrium obscures the identification of causal variants within associated loci^4^. Second, regulatory elements can influence gene expression over large genomic distances, often spanning hundreds of kilobases or even megabases^5^. Finally, gene regulation is highly cell type, developmental, and context-specific, complicating efforts to infer the biological mechanisms underlying disease associations^6^.

Expression quantitative trait locus (eQTL) mapping has emerged as a powerful approach for connecting noncoding genetic variation to gene regulation. By identifying variants associated with gene expression levels, eQTL studies when integrated with approaches such as colocalization analysis and transcriptome-wide association studies can implicate candidate target genes at GWAS loci and test whether regulatory variation mediates disease risk^7–10^. However, despite extensive eQTL mapping efforts across tissues and populations, many GWAS loci do not colocalize with currently identified eQTL signals^11,12^. For example, the GTEx Consortium found that only 43% of GWAS associations colocalized with eQTLs that were assayed across a broad range of tissues across the body^12^. This marked gap suggests that substantial disease-relevant regulatory variation remains unresolved.

Several explanations have been proposed for the limited concordance between GWAS and eQTL associations. Disease-associated variants may act in specific cell types, developmental stages, or cellular states that are not adequately represented in bulk tissue analyses^6,13^. Alternatively, differences in selective constraint, effect size distributions, or molecular mechanisms between GWAS and eQTL variants may limit detectable overlap^14^. There are also limitations to our current approaches for statistical colocalization of GWAS with eQTL associations^15^.

Standard eQTL approaches may also incompletely capture the full regulatory architecture of genes. Most eQTL studies focus on primary eQTLs, defined as the strongest regulatory association detected for a gene. However, many genes harbor multiple statistically independent regulatory variants^16–18^. These conditional or secondary eQTLs likely reflect distinct regulatory elements and may capture more context-specific regulatory programs. Recent bulk RNA-sequencing studies have shown that conditional eQTLs can colocalize with GWAS loci and reveal disease mechanisms missed by analyses restricted to primary signals^16–18^. However, the properties of genes regulated by conditional eQTLs, patterns of cell type-specificity, and disease colocalizations remain largely uncharacterized for conditional eQTLs at cell type resolution.

Here, we generated a large-scale single cell genomic resource to systematically map both primary and conditional eQTLs across major human brain cell types. By integrating single nucleus transcriptomic and whole genome sequencing data from 956 postmortem human brains and performing single cell eQTL mapping in 763 individuals of European ancestry, we identified widespread cell type-specific regulatory variation and show that conditional eQTLs represent a major and underappreciated source of genetic mechanisms underlying neuropsychiatric and neurodegenerative disease risk.

## Results

### Generation of postmortem brain single cell eQTL resource

To generate a postmortem brain single cell eQTL resource, we aggregated postmortem dorsolateral prefrontal cortex snRNA-seq data from three cohorts (**Figure 1a**)^13,19,20^. We performed rigorous harmonization to reduce batch and cohort-level effects, followed by clustering and automated cell type annotation (**Methods**)^21^.The resulting dataset comprised 4.86 million nuclei from 956 donors, similar in scale to other recently published postmortem brain snRNA-seq datasets^22,23^. Nuclei were assigned to seven major cell types: astrocytes, excitatory neurons, inhibitory neurons, endothelial cells, microglia, oligodendrocytes, and oligodendrocyte precursor cells (OPCs) (**Figure 1b; Supplementary Table S1)**. Of the 956 individuals, 529 had AD, 241 were controls, and 171 had unknown AD case-control status or other neurodegenerative diagnoses (**Figure 1a; Supplementary Table S2**).

**Figure 1:**
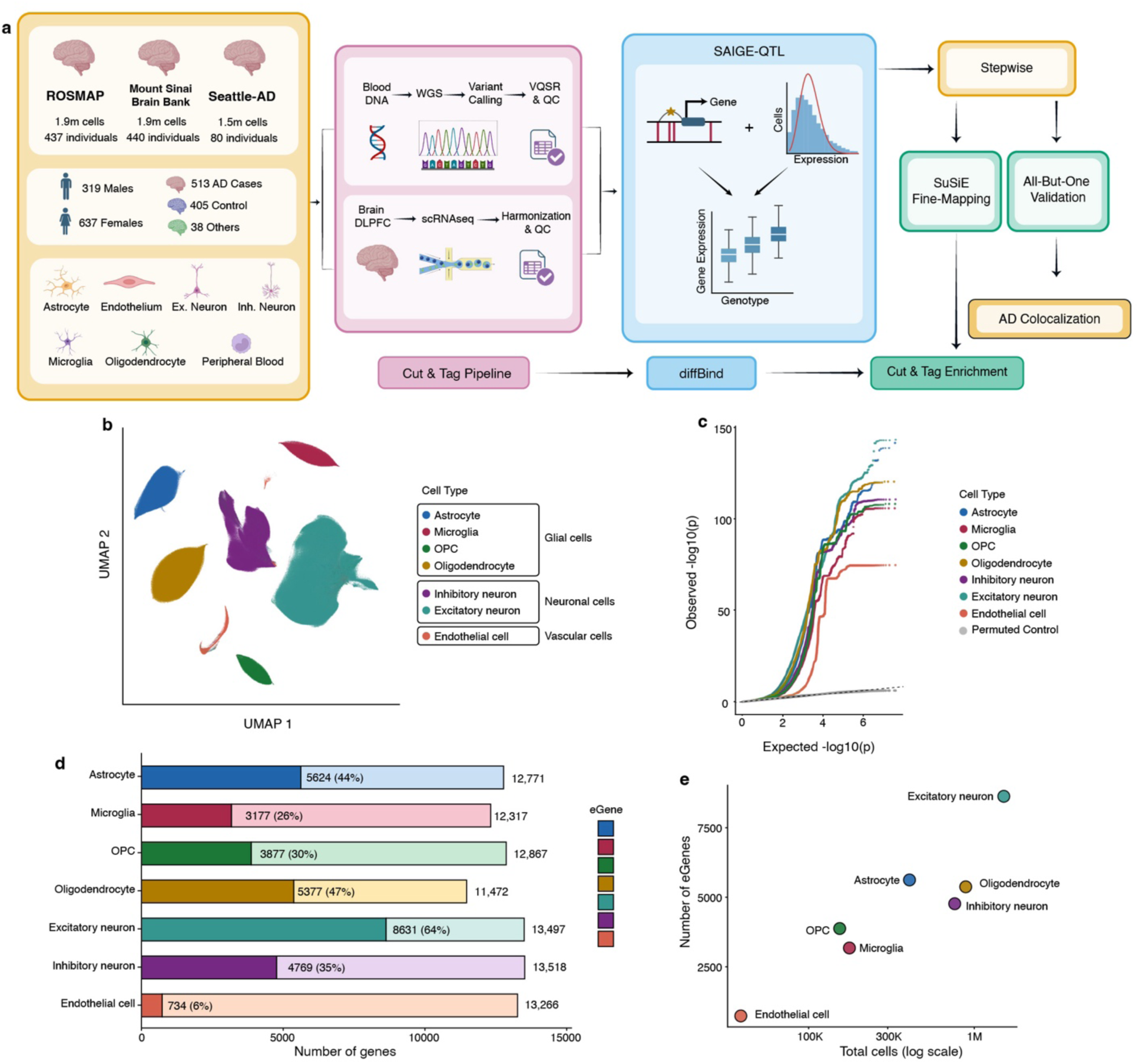
Generation of snRNA-seq resource and mapping of single cell eQTLs. a) Overview of snRNA-seq data set and workflow. b) UMAP plot of aggregated snRNA-seq dataset, colored by cell type annotations. c) QQ plot showing sc-eQTL observed p-values against expected distribution of p-values under uniform p-value distribution. Grey points show permuted null p-values across all cell types. d) Number of eGenes discovered in each cell type. Overall bar height reflects the number of testable genes in that cell type. Darker bars represent the number of significant eGenes discovered. e) Number of significant eGenes discovered in each cell type as a function of the number of cells in that cell type in the snRNA-seq dataset.

We paired gene expression data with whole genome sequencing data from each individual. To reduce sequencing batch and cohort artifacts, all samples were aligned to GRCh38 and jointly called across the full cohort, followed by rigorous quality control. We chose to perform Joint variant calling across the full cohort to improve genotyping quality and support accurate downstream statistical fine-mapping. Using principal component analysis, we retained 763 individuals of predominantly European ancestry for eQTL mapping.

We next performed cell type-specific eQTL mapping using SAIGE-QTL^24^, which models count data with a Poisson framework. Prior work has shown that modeling discrete counts can improve discovery of eQTL effects particularly in rare cell types^24,25^. We included all biallelic SNPs and indels with minor allele count >20 and tested variants within ±500 kb of each gene’s transcription start site (TSS). Association statistics were generally well calibrated, with genomic inflation factors (λ_GC_) ranging from 1.10 to 1.52 across cell types (**Figure 1c; Supplementary Table S3**). Cell types with the largest numbers of nuclei tended to show greater inflation, likely reflecting true polygenic enrichment from increased statistical power. Permutation analyses supported calibration under the null (**Figure 1c**).

We applied the aggregated Cauchy association test (ACAT), followed by genome-wide multiple testing correction using Storey’s q-value method, to identify statistically significant eGenes (**Methods**)^26^. Across cell types, we identified 734 to 8,631 eGenes per cell type (**Figure 1d**). As expected, the number of detected eGenes increased with the number of nuclei available for each cell type (**Figure 1e**).

Together, these analyses establish a large-scale resource for single cell eQTL mapping in the human brain and identify thousands of cell type-specific sc-eQTLs across major brain cell types.

### Discovery of conditional single cell eQTL effects

We next asked whether eGenes harbored additional independent regulatory effects beyond the primary sc-eQTL signal. To identify non-primary effects, we performed iterative conditional single cell *cis*-eQTL analysis followed by all-but-one validation for each sc-eQTL detected in each cell type (**Methods**) (**Figure 2a**). Across all cell types, we identified 6,265 conditional sc-eQTL signals beyond the primary association before validation (**Supplementary Table S4; Supplementary Figure S1-2**). After all-but-one validation, 6,174 conditional signals were retained, with up to seven independent sc-eQTL effects detected for a single eGene (**Figure 2b**; **Supplementary Table S5**). As expected, conditional sc-eQTL discovery increased with the number of nuclei available for each cell type.

**Figure 2:**
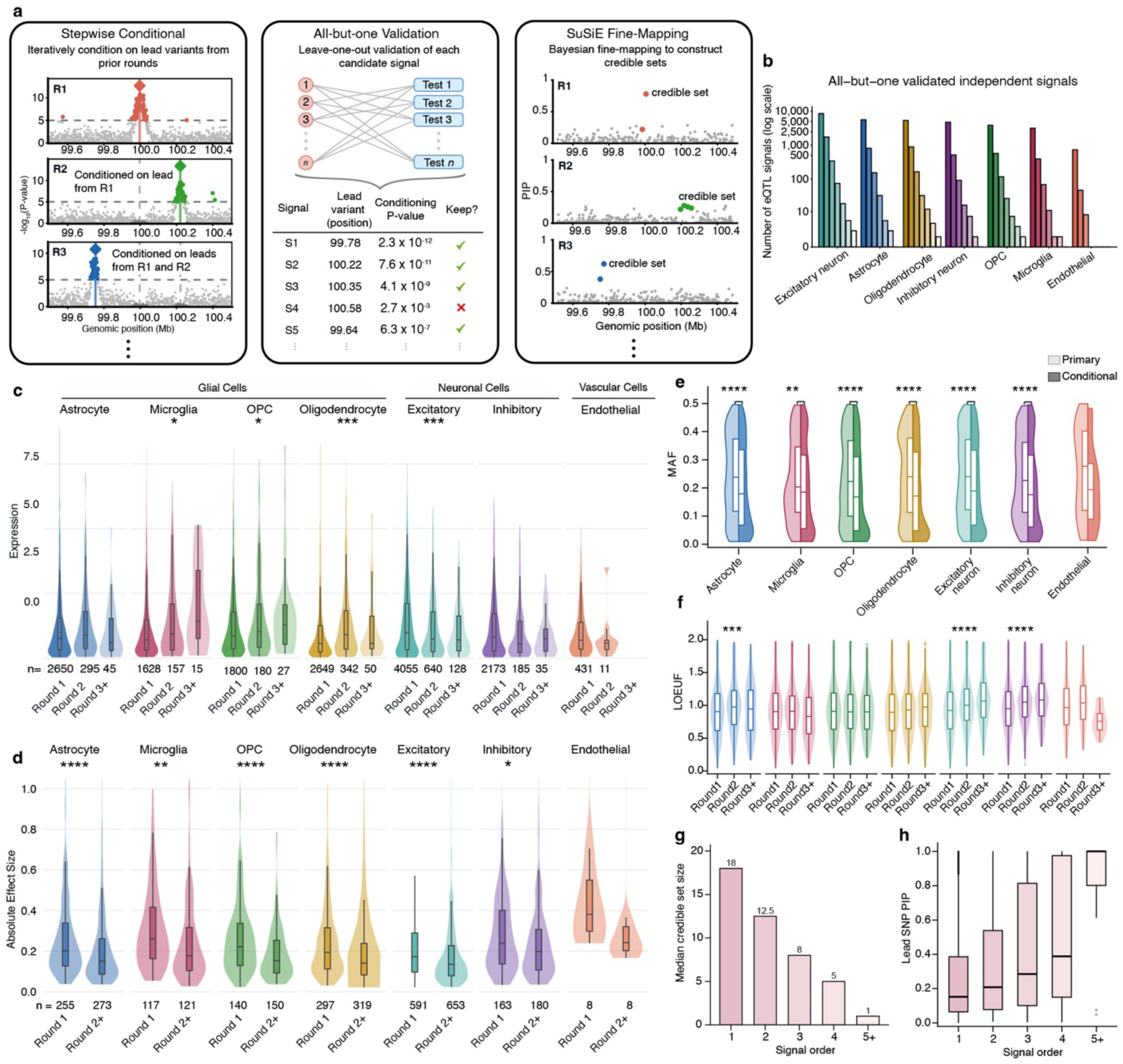
Mapping and features of conditional single cell eQTLs. a) Workflow for conditional sc-eQTL mapping. b) Number of conditional sc-eQTLs uncovered by round. Note that the y-axis is on a log_10_ scale. c) Mean expression of eGenes for round 1 primary versus round 2 conditional versus round 3+ conditional sc-eQTL signals for each cell type. Statistical significance calculated using Kruskal-Wallis test. d) Average sc-eQTL effect size for round 1 primary sc-eQTLs versus round 2+ conditional sc-eQTLs. Statistical significance calculated using Wilcoxon rank-sum test. e) MAF for lead SNP for round 1 primary sc-eQTLs versus round 2+ conditional sc-eQTLs. Statistical significance calculated using Wilcoxon rank-sum test. f) LOEUF scores of eGenes for round 1 primary versus round 2 conditional versus round 3+ conditional sc-eQTL signals for each cell type. Statistical significance calculated using Kruskal-Wallis test. g) Median fine-mapping credible set size for different rounds of sc-eQTL discovery. h) Distribution of fine-mapping PIP for most highly prioritized SNP in each round of sc-eQTL discovery. Analyses for panels d and e are restricted only to eGenes that have at least one conditional sc-eQTL in that cell type. * denotes 0.05>p≧ 0.01; ** denotes 0.01>p≧ 0.001***; denotes 0.001<p≧ 0.0001; **** denotes p<0.0001. For box plots, the center line denotes the median, box limits indicate the 25^th^ and 75^th^ percentiles, whiskers extend to values within 1.5 × IQR.

We next examined the relationship between cell type gene expression levels and the number of sc-eQTL signals detected in that cell type. For each cell type, we compared eGenes with only a primary sc-eQTL signal with eGenes with one conditional, or with two or more conditional sc-eQTLs. We found that eGenes with only a primary sc-eQTL signal showed higher expression levels than eGenes with a primary only sc-eQTL signal as compared to eGenes with one conditional, or with two or more conditional sc-eQTL signals (**Figure 2c**). Interestingly, we detected cell type-divergent effects. Excitatory neurons showed a decreased mean gene expression with additional conditional signals (p=7.2×10^-4^; Kruskal-Wallis test) and inhibitory neurons also trended to the same direction (p=0.091; Kruskal-Wallis test). In contrast, oligodendrocytes, microglia, and OPCs all showed increased mean expression in genes with additional conditional signals (p=3.2×10^-7^, 0.011, 0.024, respectively; Kruskal-Wallis tests). Together, these results show that neuronal genes with decreased expression levels tend to have more regulatory effects, while glial genes with increased expression levels tend to have more regulatory effects.

We next assessed whether primary and conditional sc-eQTLs differed in effect size and minor allele frequency distributions. For these analyses, we focused on eGenes that had at least one conditional sc-eQTL signal in a given cell type and compared the primary with conditional sc-eQTL signals for these eGenes. As expected, conditional sc-eQTLs had smaller effect sizes than their corresponding primary sc-eQTLs across all cell types (**Figure 2d**). Conditional sc-eQTLs also had lower minor allele frequencies than primary sc-eQTLs, with significant reductions observed in all cell types except endothelial cells (**Figure 2e**). Across cell types, the median minor allele frequency of conditional sc-eQTLs ranged from 0.172 to 0.194, compared with 0.207 to 0.277 for primary sc-eQTLs (**Supplementary Table S6)**. Together, these findings are consistent with reduced power to detect conditional signals, which tend to be driven by lower-frequency variants with smaller regulatory effects. Cell populations with fewer nuclei, such as endothelial cells, also showed larger effect-size distributions, consistent with reduced power to detect sc-eQTLs with smaller effect sizes.

Prior work in bulk tissues has found that eGenes with conditional eQTL signals show lower genic constraintb^18^. To test whether this pattern extends to single cell eQTLs, we integrated our data with Loss-of-function Observed/Expected Upper bound Fraction (LOEUF) scores from gnomAD (**Methods**)^27^. Lower LOEUF scores indicate stronger gene constraint, whereas higher LOEUF scores indicate reduced constraint. We found significantly higher LOEUF scores for eGenes with conditional sc-eQTL signals in astrocytes, excitatory neurons, and inhibitory neurons (**Figure 2f**; **Supplementary Table S7**). Strong differences were observed in excitatory neurons (median LOEUF 0.950 to 1.049; p=6.82×10^-19^) and inhibitory neurons (median LOEUF increasing to 1.082; p=9.28×10^-8^). We observed weaker trends in astrocytes, where median LOEUF increased from 0.904 for eGenes with only a primary sc-eQTL signal to 0.974 for eGenes with one conditional sc-eQTL signal (p=2.41×10^-4^). In contrast, no significant differences in LOEUF were observed in microglia, OPCs, oligodendrocytes, or endothelial cells. Although endothelial cells showed lower median LOEUF among eGenes with two or more conditional signals, this group contained only eight genes and the overall comparison was not statistically significant (P = 0.134). Together, these results suggest that eGenes with conditional sc-eQTL signals tend to exhibit reduced genic constraint across several cell types, with the most robust patterns observed in neuronal cell types.

We also evaluated gene constraint using the Probability of Loss-of-Function Intolerance (pLI) score from gnomAD (**Methods**)^28^, for which higher values indicate stronger genic constraint. Most cell types showed a lower proportion of highly constrained genes, defined as pLI ≥ 0.9, among eGenes with at least one conditional sc-eQTL signal compared with eGenes with only a primary sc-eQTL signal (**Supplementary Table S8**; **Supplementary Figure S3**). Microglia showed the opposite pattern: 17.3% of eGenes with only a primary sc-eQTL signal had pLI ≥ 0.9, compared with 18.9% of eGenes with at least one conditional sc-eQTL signal. These analyses indicate that eGenes with conditional signals generally have lower genic constraint across most cell types, although microglia may exhibit a distinct pattern.

Together, these results show that conditional sc-eQTLs represent a substantial layer of regulatory variation in the human brain, with thousands of independent secondary signals detected across major brain cell types. Compared with primary sc-eQTLs, conditional signals generally had smaller effect sizes, lower allele frequencies, and greater detection in higher-powered cell types, consistent with secondary regulatory effects that require large datasets to resolve. We also identified divergent patterns of constraint measures and expression levels for neuronal versus glial genes, indicating that the properties of genes regulated by more complex regulatory architectures differ across cell types.

### Statistical fine-mapping of primary and conditional sc-eQTL signals

We next performed statistical fine-mapping for each primary and conditional sc-eQTL signal using SuSiE (**Methods**) (**Figure 2a**)^29^. For each association, we used round-specific sc-eQTL summary statistics together with in-sample LD estimates derived from the whole genome sequencing data.. Primary sc-eQTL summary statistics were used to fine-map primary signals, whereas conditional summary statistics from subsequent iterations were used to fine-map secondary and higher-order conditional signals independently. Since we isolate each sc-eQTL signal through all-but-one conditional analyses, fine-mapping was performed with L = 1 for each round-specific analysis (**Methods**).

We fine-mapped 37,636 validated sc-eQTL signals across the cell types. The lead eQTL variant was contained within the resulting 95% credible set for 19,262 signals (51.6%), including 16,976 primary signals and 2,286 conditional signals. For the remaining signals, the lead variant was not retained within the credible set, suggesting greater uncertainty in causal variant localization. Credible set sizes decreased progressively with increasing signal order, with median credible set sizes of 18 variants for primary signals, 12.5 for secondary signals, 8 for tertiary signals, 5 for quaternary signals, and 1 for quinary signals (**Figure 2g**). Consistent with this pattern, posterior inclusion probabilities (PIPs) increased steadily across conditional signal order, with median PIPs of 0.153, 0.208, 0.285, 0.388, and 1.000 for primary through quinary signals, respectively (**Figure 2h**). Together, these results indicate that fine-mapping demonstrated better resolution for higher-order conditional sc-eQTL signals, despite their smaller effect sizes and lower allele frequencies.

### Conditional sc-eQTLs show more cell type-specific effects on gene expression

We first examined the extent to which validated independent sc-eQTLs and eGenes were shared across brain cell types. Most independent sc-eQTLs were detected in only one cell type (**Supplementary Figure S4**): 30,862 independent sc-eQTLs were detected in a single cell type, whereas only 14 were detected across all seven cell types (**Supplementary Figure S5a**). At the gene level, eGenes showed broader sharing across cell types, although 3,830 eGenes were detected in only one cell type (**Supplementary Figure S5b**). To determine whether cell type-specific eGene discovery reflected cell type-specific expression, we examined the breadth of expression of eGenes detected in only one cell type. Among single cell type eGenes, 1,951 were expressed in all seven cell types, whereas only 466 were expressed in a single cell type (**Supplementary Figure S5c-d**). These results suggest that cell type-specific sc-eQTL discovery is not explained solely by cell type-restricted gene expression patterns.

We also examined whether individual sc-eQTLs were associated with multiple target genes in a given cell type. Most independent sc-eQTLs across all cell types were associated with a single eGene, whereas multi-gene regulatory effects were uncommon and usually limited to two target genes (**Supplementary Figure S6a-b**). This pattern was observed for both primary and conditional sc-eQTLs, but was particularly pronounced for conditional signals. Among primary sc-eQTLs with multiple target genes, 411 were associated with two eGenes and 48 were associated with three or more eGenes (**Supplementary Figure S6c**). In contrast, only 71 conditional sc-eQTLs were associated with more than one eGene, and most of these regulated expression of only two eGenes (58 of 71, 81.7%) (**Supplementary Figure S6d**). These results suggest that conditional sc-eQTLs are less likely to have broad multi-gene regulatory effects and instead tend to act in a target gene-specific manner.

We next tested whether primary and conditional eQTL signals differed in their sharing across cell types. We applied mashr, a Bayesian approach for effect-size sharing analysis^30^. For each pairwise cell type comparison, we analyzed significant SNP-gene pairs detected in both cell types and estimated sharing of effect direction and effect magnitude separately for primary and conditional single cell eQTL signals (**Methods**). Across cell types, effect direction sharing was consistently high for both primary and conditional sc-eQTLs, with sign-sharing generally ranging from 0.82 to 0.96 across cell type pairs (**Supplementary Figure S7**). In contrast, magnitude sharing was more variable, ranging from 0.46 to 0.80 for primary sc-eQTLs and from 0.35 to 0.75 for conditional sc-eQTLs (**Figure 3a**). Overall, conditional sc-eQTLs generally exhibited reduced effect magnitude sharing across pairs of cell types, consistent with greater cell type specificity of secondary regulatory effects. Endothelial cell pairs showed relatively high estimated sharing for conditional sc-eQTLs, although these estimates were based on fewer overlapping significant SNP-gene pairs than most other cell type pairs (n = 460-599 overlapping SNP-gene pairs for endothelial pairwise comparisons) (**Supplementary Table S9**).

**Figure 3:**
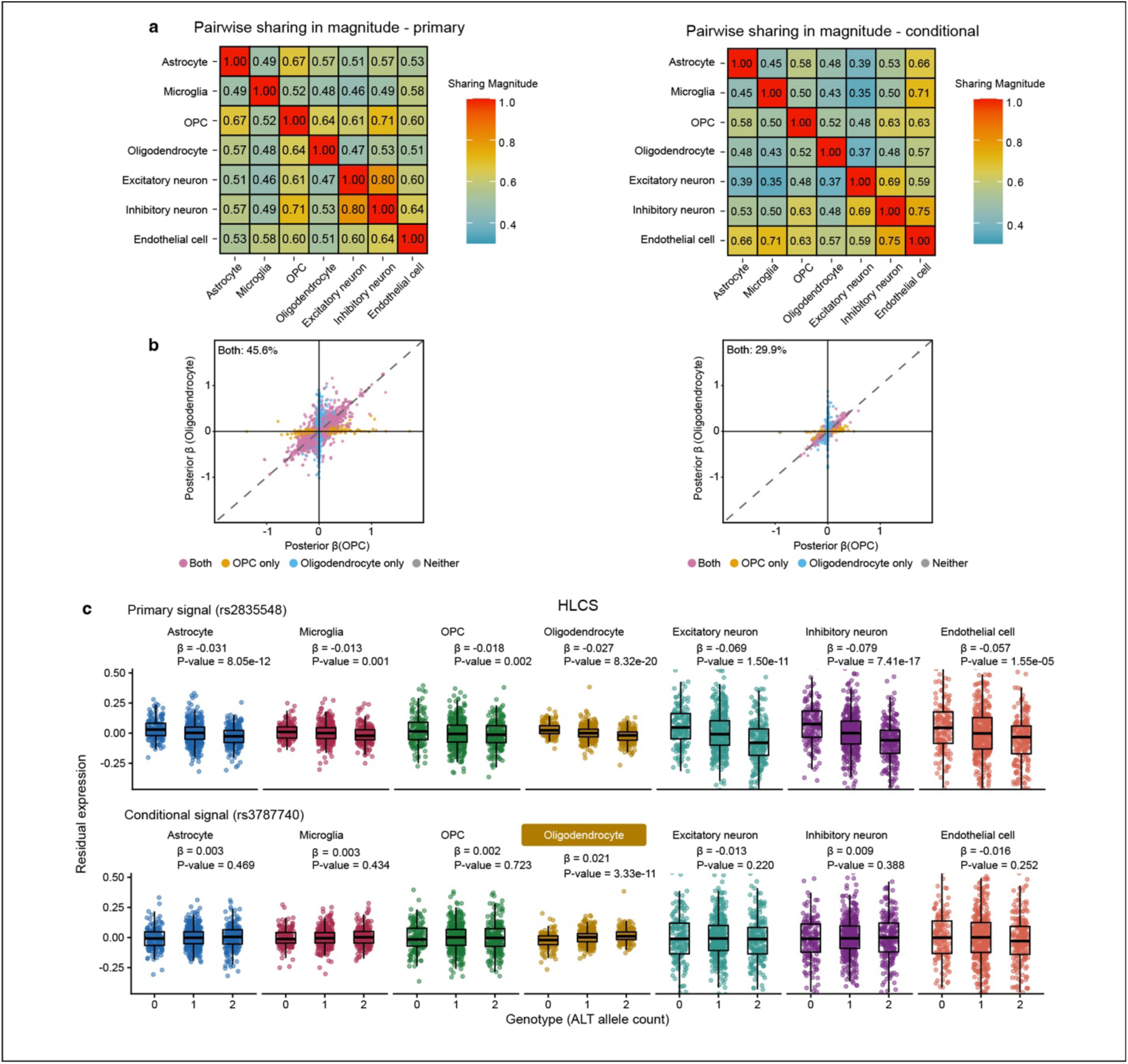
Cell type specificity of single cell eQTLs. a) Pairwise effect magnitude sharing of sc-eQTL signals for primary (left) versus secondary (right) sc-eQTLs. b) Comparison of posterior β values for oligodendrocytes versus OPCs from pairwise mashr test. Each point represents one SNP-gene pair. Points are colored by significance status: both cell types are significant (pink), oligodendrocytes only (yellow), OPCs only (blue), or neither (gray). c) Violin plots of *HCLS* expression across cell types for primary sc-eQTL signal (rs2835548, top) and conditional signal (rs3787740, bottom). Each point is the mean *HCLS* expression for an individual in that cell type.

For example, pairwise mashr analysis demonstrated that oligodendrocytes and OPCs shared 64% of primary eQTL effects in effect magnitude, compared with 52% of conditional eQTL effects (**Figure 3b**). Consistent with this pattern, direct comparison of posterior effect sizes showed that 45.6% of primary eQTL pairs were significant in both cell types, whereas only 29.9% of conditional eQTL pairs were significant in both cell types (**Figure 3b**).

For the gene *HLCS*, the primary signal (rs2835548) showed statistically significant and directionally concordant sc-eQTL effects across oligodendrocytes and OPCs (**Figure 3c**). In contrast, the conditional signal (rs3787740) showed a statistically significant sc-eQTL effect only in oligodendrocytes (p=3.33×10^-11^; β=0.021). Together, these results demonstrate that conditional eQTLs reflect more cell type-specific genetic effects on gene expression.

### Epigenetic overlap of primary and conditional sc-eQTLs

We next asked whether primary and conditional sc-eQTLs differed in their noncoding regulatory features. We first compared the distance of primary and conditional sc-eQTLs to the TSS of the regulated gene, considering only eGenes with at least one conditional sc-eQTL in a given cell type. Conditional sc-eQTLs were systematically farther from the TSS than primary sc-eQTLs (**Figure 4a**), a pattern observed across all cell types (**Supplementary Figure S8**). When all eGenes were considered, regardless of whether they had a conditional sc-eQTL, distance to the TSS increased with successive conditional sc-eQTL rounds (**Supplemental Figure S9**). These patterns were also reflected in reduced promoter-proximal annotation and increased distal intergenic annotation among conditional sc-eQTLs compared with primary sc-eQTLs (**Figure 4b; Supplemental Figure S10**).

**Figure 4:**
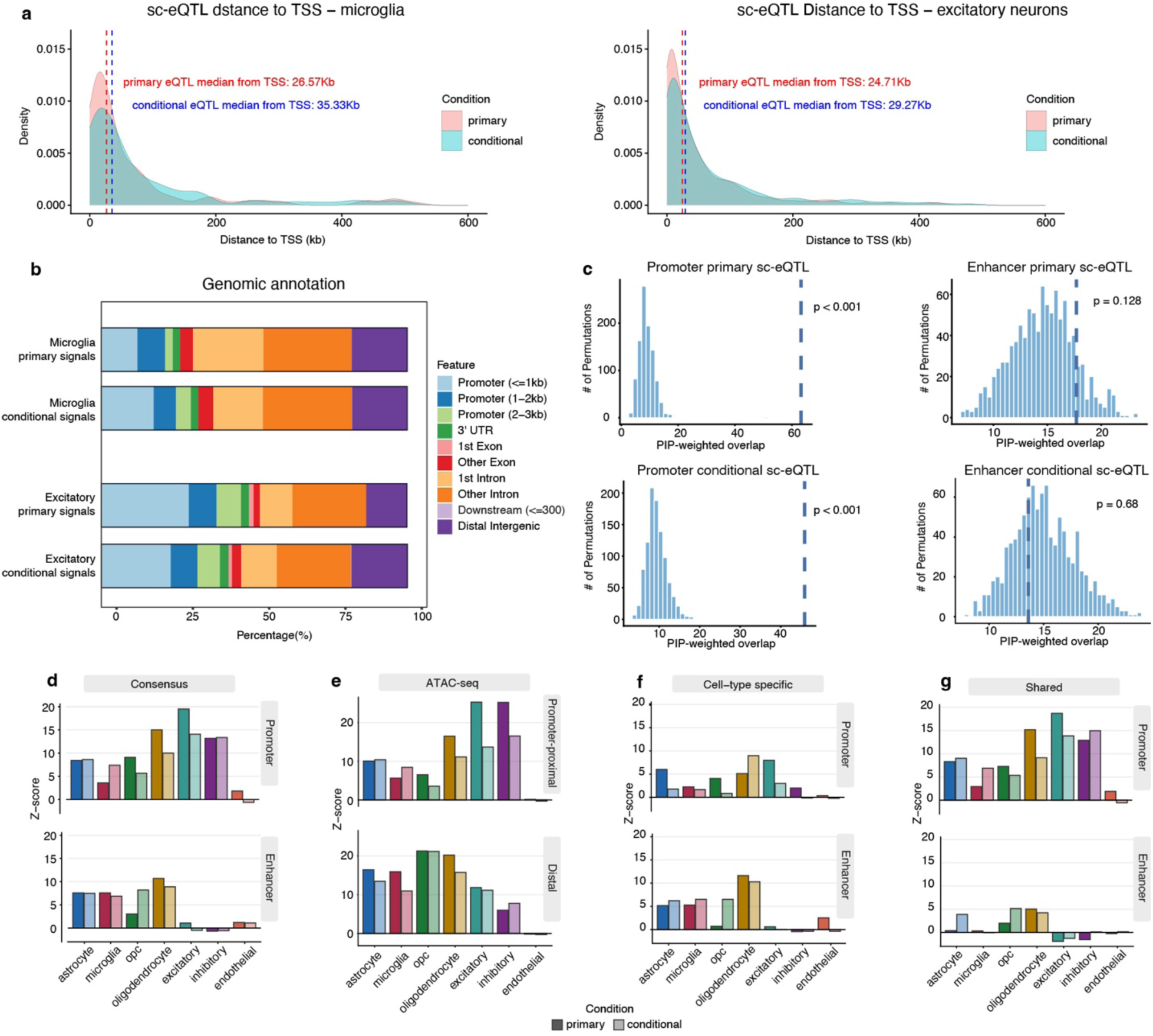
Epigenetic enrichments of single cell eQTLs. a) Distance to TSS of cognate eGene for the lead SNP of primary versus conditional sc-eQTLs in microglia (left) and excitatory neurons (right). b) Genomic distributions of lead SNP of primary versus conditional sc-eQTLs in microglia (top) and excitatory neurons (bottom). c) Histograms of random permutations of epigenetic overlaps in excitatory neurons. Histogram shows distribution of PIP-weighted overlaps from 1000 permutations. Dotted vertical line shows observed PIP-weighted overlap. Separate histograms are shown for consensus promoter and enhancer regions in excitatory neurons, each for primary and conditional sc-eQTLs. d) Z-score of permutation analysis for consensus promoters and enhancers as defined by H3K4me3 and H3K27ac CUT&Tag data. Results are stratified by cell type and primary versus conditional. e) Same as d), except using promoter-proximal OCRs and distal OCRs from snATAC-seq data. f) Same as d), except for cell type-specific CUT&Tag promoters and enhancers. g) Same as d), except for cell type-shared CUT&Tag promoters and enhancers. For all panels, results are only shown for eGenes that have at least one conditional sc-eQTL in that cell type.

To assess the regulatory context of these eQTLs, we obtained and reprocessed bulk CUT&Tag data from sorted postmortem brain cell populations^31^. We analyzed H3K4me3 which marks active promoters and H3K27ac which marks active promoters and enhancers. We identified peaks for each histone mark and cell type, including putative promoter peaks (H3K4me3-positive peaks within 3 kb of a TSS) and putative enhancer peaks (H3K27ac-positive, H3K4me3-negative peaks located more than 3 kb from a TSS). For each cell type, we also classified promoter and enhancer peaks as cell type-shared or cell type-specific (**Methods**).

We next tested whether sc-eQTL signals were enriched in regulatory regions from each cell type and whether these enrichments differed between primary and conditional signals. We restricted this analysis to eGenes with at least one conditional sc-eQTL signal in the relevant cell type. For each credible set, we calculated observed overlap between sc-eQTL variants and epigenetic peaks, weighted by fine-mapping PIP (**Methods**). To assess significance, we used a GoShifter-inspired permutation strategy^32^. In this approach, epigenetic annotations are randomly shifted within a circularized 1-Mb *cis-*window centered on the TSS (±500 kb), and PIP-weighted overlaps are recalculated for each permutation. We then computed a Z-score by subtracting the mean permuted PIP-weighted overlap from the observed weighted overlap and dividing by the standard deviation of the permuted weighted overlaps (**Figure 4c**). This approach is less sensitive to biases from non-random distributions of local genomic annotations^32^ and avoids potential biases from matching random SNPs, which is particularly important given the allele-frequency differences between primary and conditional signals.

Among genes with at least one conditional sc-eQTL signal in a given cell type, both primary and conditional sc-eQTLs were strongly enriched in active promoter regions (**Figure 4d**). In most cell types, primary signals were more enriched in active promoters than corresponding conditional signals. Microglia were the main exception, showing stronger promoter enrichment for conditional signals than for primary signals (**Figure 4d**). We also examined putative enhancer regions. Overall enrichment in putative enhancers was lower than enrichment in promoter regions across cell types (**Figure 4d**), consistent with prior observations^14^. This pattern was especially pronounced in excitatory and inhibitory neurons, where putative enhancer enrichment was limited. For most cell types, primary and conditional signals showed similar enhancer enrichment, although OPCs showed stronger enrichment for conditional sc-eQTL signals than for primary signals.

We performed analogous analyses using open chromatin regions (OCRs) called from snATAC-seq data^33^. OCR peaks were previously generated from pseudobulked cell populations in this dataset. We partitioned OCRs into promoter-proximal peaks (≤3 kb from a TSS) and distal peaks (>3 kb from a TSS). Similar to the histone CUT&Tag analyses, we observed strong enrichment in promoter-proximal OCRs across cell types, with generally stronger enrichment for primary than conditional sc-eQTL signals; microglia again were an exception. In contrast to the histone CUT&Tag results, glial cell types (including astrocytes, microglia, oligodendrocytes, and OPCs) showed stronger enrichment in distal OCRs than in promoter-proximal OCRs. These divergent results suggest that distal OCRs may capture regulatory effects of eQTL variants not represented by canonical histone mark-based enhancer annotations, particularly in glial cell types (**Figure 4e**).

Returning to the CUT&Tag data, we next partitioned promoters and putative enhancers into cell type-shared and cell type-specific elements (**Methods**). Overall, sc-eQTLs showed stronger enrichment in cell type-shared promoters and weaker enrichment in cell type-shared putative enhancers (**Figure 4f-g**). Neuronal cell types were enriched almost exclusively in cell type-shared promoters, with limited or no enrichment in cell type-specific promoters or cell type-shared putative enhancers.

Together, these results show that sc-eQTLs are most consistently enriched in cell type-shared promoter regions. They also reveal strong enrichment in distal OCRs, suggesting that chromatin accessibility can capture regulatory elements not represented by canonical histone-based enhancer annotations.

### Colocalization of primary and conditional sc-eQTL signals with disease-associated GWAS loci

To determine whether primary and conditional sc-eQTL signals colocalize with disease-associated loci, we performed Bayesian colocalization analysis using GWAS summary statistics for Alzheimer’s disease (AD) from Bellenguez et al. and schizophrenia (SCZ) from Trubetskoy et al. (**Figure 5a**) (**Methods**)^34,35^. Across all cell types, we conducted 30,315 sc-eQTL signal-level colocalization tests across 10,312 genes for AD and 30,308 tests across 10,311 genes for SCZ^36^. Using a PP.H4 threshold of 0.5, we identified 373 AD and 867 SCZ colocalization signals, including 104 and 151 high-confidence colocalizations (PP.H4>0.9), respectively (**Supplementary Table S10**). Excitatory neurons and oligodendrocytes showed the greatest number of SCZ colocalization signals, whereas AD colocalization signals were more broadly distributed across cell types (**Supplementary Figure S11**).

**Figure 5:**
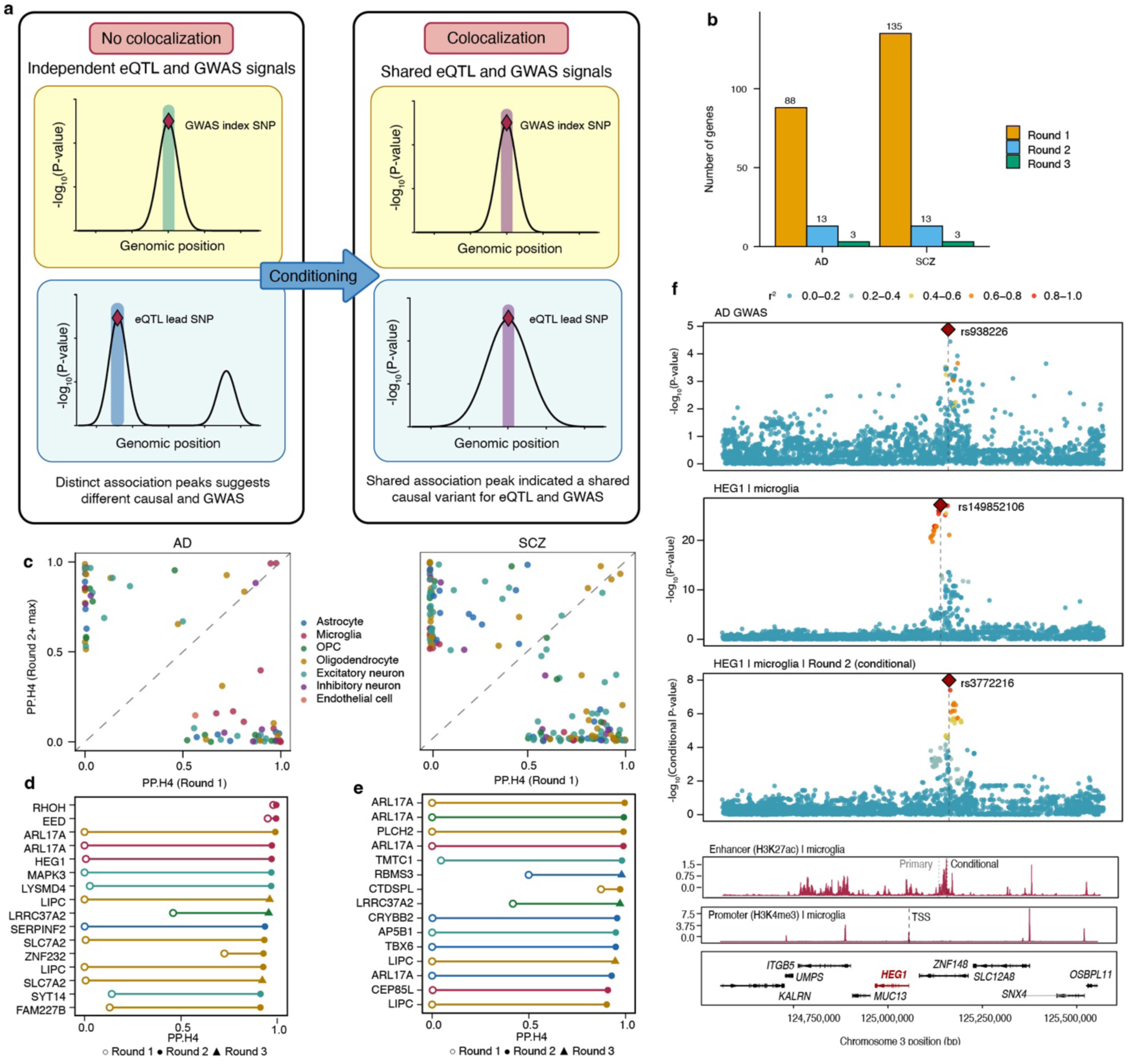
Colocalization of single cell eQTLs with AD and SCZ GWAS. a) Conceptual overview for colocalization with GWAS. b) Number of eGenes colocalizing with AD and SCZ GWAS by conditional round. c) Comparison of PP.H4 between primary and conditional sc-eQTL signals. Points above the diagonal indicate loci where the conditional signal showed stronger colocalization evidence than the primary signal. d, e) AD (d) and SCZ (e) examples where conditional sc-eQTL signals showed stronger colocalization evidence than the corresponding primary signal. f) Locuszoom plot showing AD GWAS results at *HEG1* locus (top panel), marginal sc-eQTL results for *HEG1* in microglia (middle panel), and round 2 conditional sc-eQTL results for *HEG1* in microglia (bottom panel). Locuszoom plots are overlaid on microglia H3K27ac and H3K4me3 CUT&Tag data.

Most colocalization signals originated from primary sc-eQTL associations. Among signals with PP.H4 >0.5, 329 of 373 AD colocalizations and 780 of 867 SCZ colocalizations corresponded to primary sc-eQTL signals (**Figure 5b; Supplementary Table S10**). However, conditional sc-eQTL signals also contributed to disease colocalization. For AD, 44 conditional sc-eQTL signals exceeded PP.H4>0.5, including 35 secondary, 8 tertiary, and 1 quaternary signal. For SCZ, 87 conditional sc-eQTL signals exceeded PP.H4>0.5, including 76 secondary, 9 tertiary, and 2 quaternary signals (**Supplementary Table S10**). At the high-confidence threshold of PP.H4 >0.9, we identified 16 colocalizations for conditional sc-eQTLs for AD and 16 for SCZ, including 13 secondary and 3 tertiary signals for each disease.

We next examined whether conditional sc-eQTL signals showed stronger colocalization than the corresponding primary sc-eQTL signals at the same locus. Among conditional colocalizations with PP.H4 >0.5, 42 of 44 AD signals (95.5%) and 82 of 87 SCZ signals (94.3%) had higher PP.H4 values than their corresponding primary signals (**Figure 5c**). Similar patterns were observed for high-confidence colocalizations (PP.H4 >0.9), where all 16 AD conditional signals and 15/16 SCZ conditional signals showed stronger colocalization evidence than the corresponding primary signal (**Figure 5d-e**). These results suggest that although most disease colocalizations arise from primary sc-eQTL signals given the sheer number of primary signals, in many instances, conditional sc-eQTLs can provide stronger evidence for a shared causal variant as compared to a primary sc-eQTL signal.

As an example, we examined the *HEG1* locus in microglia, where a conditional sc-eQTL signal showed strong colocalization with AD GWAS (**Figure 5f**). At this locus, the AD GWAS association and primary *HEG1* sc-eQTL association were not well aligned (PP.H4=0.007). After conditioning on the primary sc-eQTL signal, a secondary microglia *HEG1* signal emerged that overlapped the AD GWAS peak and was far from the TSS (PP.H4=0.97). This conditional *HEG1* signal remained significant after conditioning (p=1.01×10^-8^; β=-0.17) and mapped to a microglia enhancer-marked region, supporting its potential relevance to microglial gene regulation (**Figure 5f**).

Together, these results demonstrate that although most disease-associated colocalizations involve primary sc-eQTL signals, conditional sc-eQTLs often provide the strongest evidence for a shared causal variant and can reveal disease-relevant regulatory mechanisms that would be overlooked when considering only the primary signal.

## Discussion

In this study, we generated a large-scale human post-mortem brain single cell eQTL resource and used it to map both primary and conditional regulatory effects across major brain cell types. By performing single cell eQTL analyses in 763 individuals of European ancestry across genes from seven major brain cell types, we identified 32,189 eGenes and 6,174 additional independent conditional sc-eQTL signals. These analyses reveal that conditional sc-eQTLs represent a substantial layer of regulatory variation in the human brain. Compared with primary sc-eQTLs, conditional signals were generally more cell type-specific, located farther from TSSs, and less enriched in canonical promoter and enhancer annotations. Genes harboring conditional sc-eQTLs also showed distinct patterns of relaxed selective constraint, particularly in neuronal cell types. Finally, colocalization analyses with AD and SCZ GWAS demonstrated that conditional sc-eQTLs can provide stronger evidence for shared causal variation than primary sc-eQTLs at some disease-associated loci. Together, these findings show that conditional regulatory effects provide additional resolution for interpreting the genetic architecture of brain traits and diseases.

A major finding of this work is that conditional sc-eQTLs are widespread in the human brain and capture regulatory effects that are not apparent from primary sc-eQTL analyses alone. Most eQTL studies focus on the strongest association for each gene, yet many genes are regulated by multiple independent variants. Our results extend prior work from bulk tissues by showing that this multi-signal regulatory architecture is readily detectable in single cell data when sample size and cell counts are sufficiently large^16–18^. Conditional sc-eQTLs tended to have smaller effect sizes and lower allele frequencies than primary sc-eQTLs, consistent with the expectation that secondary regulatory effects are more difficult to detect and require greater statistical power. These findings underscore the value of large-scale single cell datasets for resolving regulatory architectures that are likely missed in smaller studies.

We also found that genes with conditional sc-eQTLs often differed in selective constraint from genes with only primary sc-eQTLs. Across several cell types, and most prominently in neurons, eGenes with conditional signals showed higher LOEUF scores and a lower proportion of high-pLI genes, consistent with reduced selective constraint. This pattern is broadly concordant with prior analyses in bulk tissues, which found that eGenes with conditional eQTLs tend to be less constrained^18^. One possible interpretation is that genes under weaker constraint may tolerate a greater number of common regulatory variants, making multi-signal architectures more detectable. Alternatively, reduced constraint may reflect differences in gene function, expression breadth across cell types/tissues, or regulatory architectures that are only partially captured by the presence of conditional eQTLs. Notably, these patterns were not uniform across all cell types, and microglia showed a different trend in some analyses. Thus, while conditional sc-eQTLs appear to be linked to differences in gene-level constraint, the relationship between regulatory complexity and selection likely differs across brain cell classes.

Conditional sc-eQTLs also showed evidence of greater cell type-specificity than primary sc-eQTLs. Although many eGenes were expressed across multiple cell types, their associated eQTLs were often detected in only one or a subset of cell types, indicating that cell type-specific eQTL discovery is not simply explained by cell type-restricted expression. Pairwise mashr analyses further showed that primary and conditional sc-eQTLs frequently shared effect direction across cell types, but conditional signals generally showed weaker sharing of effect magnitude. This suggests that secondary regulatory variants often influence expression of the same target genes across related cell types, but with more context-dependent effect sizes. Such quantitative differences may be especially important in the brain, where shared genes can be embedded in distinct chromatin landscapes, transcriptional programs, and cellular states across neurons and glia.

Our epigenetic analyses further suggest that primary and conditional sc-eQTLs differ in their regulatory context. Conditional sc-eQTLs were systematically farther from the TSSs of their target genes than primary sc-eQTLs, consistent with prior findings in bulk eQTL studies^17,18^. We initially expected that this increased TSS distance would correspond to stronger enrichment of conditional sc-eQTLs in enhancer annotations. However, conditional signals were not more enriched in histone-marked enhancers. Instead, both primary and conditional sc-eQTLs showed the strongest enrichment in promoter-proximal regulatory regions, and conditional signals generally showed weaker enrichment in both promoters and enhancers. These findings suggest that conditional sc-eQTLs are not simply a distal enhancer-enriched class of regulatory variants. We note that Brotman et al had similar findings in adipose bulk eQTL, where each round of conditional eQTLs had progressively decreased enrichments in promoters and enhancers^18^. Instead, conditional sc-eQTLs may reflect a broader set of regulatory mechanisms, some of which are not well-captured by canonical H3K27ac/H3K4me3-based annotations. Comparison with snATAC-seq data provided an additional layer of insight. In glial cell types, sc-eQTLs showed stronger enrichment in distal OCRs than in histone-defined enhancer annotations. This divergence suggests that distal OCRs may capture regulatory elements or chromatin contexts not fully represented by canonical active histone marks, particularly in glial cells. These results also have practical implications for variant prioritization: approaches that prioritize noncoding eQTLs solely based on promoter or histone-marked enhancer overlap may miss regulatory variants located in other accessible chromatin contexts.

Several results point to differences in regulatory architecture between neuronal and glial cell types. Neuronal eGenes with conditional sc-eQTLs tended to have lower expression and reduced constraint. Neuronal sc-eQTLs were strongly enriched in cell type-shared promoter regions, with limited enrichment in enhancer annotations. In contrast, glial genes with conditional sc-eQTLs tended to have higher expression levels in eGenes with conditional sc-eQTL signals and little difference in measures of constraint. Glial sc-eQTLs also showed more balanced enrichment across promoters and enhancers, including stronger enrichment in cell type-specific enhancer elements. These observations suggest that conditional sc-eQTLs may capture different forms of regulatory complexity across brain cell classes. In glia, conditional signals may reflect additional enhancer-mediated inputs into genes that are already robustly expressed within lineage or state-specific regulatory programs. In neurons, conditional signals may be more detectable at lower-expression, less constrained genes where additional common regulatory variation is more tolerated. These interpretations remain speculative, but they highlight how conditional single cell eQTL mapping can reveal cell class-specific differences in the relationship between gene expression, chromatin context, and selective constraint.

A key motivation for mapping conditional sc-eQTLs is to improve interpretation of disease-associated noncoding variation. As expected, with AD and SCZ GWAS most colocalized signals involved primary sc-eQTLs. However, we also identified multiple secondary and higher-order conditional sc-eQTLs with strong colocalization evidence. Importantly, when conditional sc-eQTLs colocalized with disease-associated loci, they often showed stronger colocalization probabilities than the corresponding primary sc-eQTL signal for the same gene. This indicates that primary eQTLs can obscure disease-relevant secondary regulatory effects at some loci. Conditional sc-eQTL mapping therefore provides a complementary strategy for resolving GWAS loci.

This study has several limitations. First, although our dataset is large, analyses were restricted to broad major brain cell types. More granular cell subclasses, such as cortical projection neuron subtypes or disease-associated microglial states, may harbor additional context-specific regulatory effects that could not be robustly detected here. Second, differences in cell counts across cell types influence power to detect both primary and conditional sc-eQTLs, limiting quantitative comparisons across cell populations. We therefore interpret cross-cell type differences qualitatively rather than as definitive rankings of regulatory complexity. Third, fine-mapping was performed using summary statistics from count-based SAIGE-QTL models, whereas SuSiE assumes a linear regression framework as has been discussed previously^24,25^.

In summary, our study provides a large-scale map of primary and conditional sc-eQTLs across major human brain cell types. These data show that conditional regulatory effects are widespread, often more cell type specific than primary effects, and can reveal disease-relevant regulatory mechanisms that may be missed by primary eQTL analyses alone. More broadly, our findings highlight the importance of modeling multi-signal regulatory architecture in single cell datasets to better understand how noncoding genetic variation shapes gene regulation in the human brain and contributes to neuropsychiatric and neurodegenerative disease risk.

## Methods

### Ethics statement

This study is based exclusively on the analysis of publicly available human genetic data. All contributing studies received ethical approval from their respective institutional review boards, and informed consent was obtained from all participants, as described in the original publications.

### snRNA-seq data integration and harmonization

We assembled a snRNA-seq dataset from three publicly available cohorts obtained through Synapse: ROSMAP (syn3219045), SEA-AD (syn26223298), and MSSM PsychAD (syn52160016)^19,20,35^. Raw count matrices were downloaded from each cohort, converted to AnnData format, merged, and processed using Scanpy v1.12^37^ and RAPIDS-singlecell (v0.14.1; https://rapids-singlecell.readthedocs.io/en/latest/).

Quality control was performed prior to normalization and integration. Doublets were identified and removed using Scrublet, as implemented in sc.pp.scrublet. Nuclei with >15% mitochondrial reads were excluded. We also removed nuclei for which either ln(1 + total counts) or ln(1 + detected genes) deviated by more than four median absolute deviations from the dataset median. Counts were then normalized using rapids_singlecell.pp.normalize_total, log-transformed with log1p, restricted to the top 2,000 highly variable genes, and z-score scaled using RAPIDS-singlecell functions.

Dimensionality reduction and integration were performed on the processed expression matrix. Principal components were computed with rapids_singlecell.pp.pca using 50 components, and batch effects were corrected using Harmony as implemented in rapids_singlecell.pp.harmony_integrate^38^. A nearest-neighbor graph was then constructed using 40 neighbors and 40 principal components. This graph was used for UMAP visualization and Leiden clustering at resolution 0.4.

Cell types were initially annotated using CellTypist v1.7.1 with the preloaded adult human prefrontal cortex reference model^21^. To identify poorly resolved or ambiguous clusters, we calculated cluster marker genes using sc.tl.rank_genes_groups and scored canonical cell type marker sets using sc.tl.score_genes. Clusters were removed if they expressed markers from contradictory cell types or lacked a clearly dominant canonical marker signature. The final dataset was restricted to donors of European ancestry with paired whole genome sequencing data.

### WGS alignment, variant calling and QC

Whole genome sequencing FASTQ files were obtained from NIAGADS (https://dss.niagads.org/datasets/ng00067/ and https://dss.niagads.org/datasets/ng00174/). Per-sample paired-end fastq files were aligned to the GRCh38 reference genome (GCA_000001405.15, no-alt analysis set), and germline variants were called using the NVIDIA Clara Parabricks germline pipeline v4.5.1-1 (pbrun germline). This pipeline provides a GPU-accelerated implementation of the GATK Best Practices workflow for germline variant discovery^39–41^.

For each sample, reads were aligned with bwa-mem using -K 10000000^42^. The resulting alignments were coordinate-sorted, duplicate-marked, and subjected to base quality score recalibration (BQSR). BQSR was performed using three known-variant resources: Mills and 1000 Genomes gold-standard indels for hg38, the GATK Homo_sapiens_assembly38 known-indels resource, and dbSNP build 146 for hg38. Recalibrated alignments were then processed with HaplotypeCaller in GVCF mode to generate per-sample genomic VCFs (.g.vcf.gz) for downstream joint genotyping. Sorting and BAM writing were accelerated using GPU-enabled options in Parabricks (--gpusort, --gpuwrite).

Per-sample gVCFs were aggregated into a GATK GenomicsDB workspace using GATK v4.1.7.0 and jointly genotyped^40,41^. The resulting cohort-level VCFs were filtered according to GATK Best Practices for germline short-variant discovery. Variants with ExcessHet >54.69 were flagged using VariantFiltration, and a sites-only VCF was generated with MakeSitesOnlyVcf for Variant Quality Score Recalibration (VQSR) model training.

VQSR was performed separately for indels and SNPs. Indel recalibration with --max-gaussians 4 and was trained on Mills and 1000 Genomes gold-standard indels, Axiom Exome Plus, and dbSNP 138. SNP recalibration with --max-gaussians 6 was trained on HapMap 3.3, 1000 Genomes Omni 2.5, 1000 Genomes Phase 1 high-confidence SNPs, and dbSNP 138. For both variant classes, we used the resource priors and annotation sets recommended by GATK Best Practices^41^. Recalibration was applied with ApplyVQSR using truth-sensitivity thresholds of 99.7% for indels and 99.5% for SNPs. The recalibrated SNP and indel callsets were then combined with MergeVcfs. Variant calling metrics were summarized against dbSNP 138 using CollectVariantCallingMetrics.

Following joint genotyping and VQSR, variants were further processed using PLINK 2.0^43^. The recalibrated callset was restricted to autosomal chromosomes and European-ancestry individuals. Hardy-Weinberg equilibrium (HWE) filtering was performed in controls only, excluding variants with HWE p < 1 × 10⁻⁶. The HWE-passing variant list was then used to subset the dataset before applying additional quality-control filters. We removed variants with genotype missingness >10% (--geno 0.1), minor allele frequency <1% (--maf 0.01), or minor allele count <20 (--mac 20). Samples with per-individual missingness >10% were excluded (--mind 0.10), and only biallelic variants were retained (--max-alleles 2).

### PEER factor selection

Prior to PEER factor estimation, gene expression was aggregated to the donor level within each cell type by averaging single-nucleus counts, generating a cell type-specific pseudo-bulk expression matrix. Genes with zero expression in ≥90% of donors within a given cell type were excluded. The remaining expression values were log-normalized using sc.pp.log1p, and the top 2,000 highly variable genes were selected based on dispersion using sc.pp.highly_variable_genes with n_top_genes=2,000. Expression values were then scaled to zero mean and unit variance using sc.pp.scale.

PEER factors were estimated separately for each cell type using peertool v1.3^44^. For each cell type, 50 PEER factors were inferred while including age, sex, postmortem interval, the first 10 genotype principal components, and dataset of origin as covariates.

### sc-eQTL mapping using SAIGE-QTL

For each cell type, genes detected in fewer than 1% of nuclei were excluded from SAIGE-QTL input to remove lowly expressed features. Genotype data were pruned to obtain a set of approximately independent SNPs for model fitting and variance-ratio estimation. LD pruning was performed with PLINK 2.0 using a sliding window of 50 variants, a step size of 5 variants, and an r² threshold of 0.2 (--indep-pairwise 50 5 0.2). A reduced genotype dataset was then generated from the pruned SNP list. To minimize computational burden while retaining representative genome-wide variation, we randomly selected 2,000 SNPs from the LD-pruned set for use in SAIGE-QTL model fitting.

Single cell *cis*-eQTL mapping was performed using SAIGE-QTL^24^. For each gene within each cell type, we fit a null generalized linear mixed model using step1_fitNULLGLMM_qtl.R with raw UMI counts as the input (--traitType=count). The log-transformed total UMI count per nucleus was included as an offset to account for differences in sequencing depth across nuclei (--offsetCol=log1p_total_counts). Variance-ratio estimation was performed using the random subset of 2,000 LD-pruned SNPs described above.

Fixed-effect covariates included postmortem interval, age at death, sex, dataset of origin, PEER factors, mitochondrial read fraction, and genotype principal components. We included genotype PCs 1, 2, 4, and 5; PC3 was excluded because it was strongly collinear with SEA-AD dataset membership. Covariate transformation was applied before model fitting (--isCovariateTransform=TRUE), and zero-count observations were retained in the phenotype vector (--isRemoveZerosinPheno=FALSE). Models were fit using a convergence tolerance of 0.001, a maximum of 50 outer iterations, and a maximum of 5,000 inner conjugate-gradient iterations (--maxiterPCG=5000).

To identify genes with residual model misspecification or inflated null association statistics, we performed a permutation-based calibration analysis. Donor IDs in the genotype file were shuffled, and the SAIGE-QTL null model was tested on the permuted data. QQ plots were generated for each cell type to assess the empirical null distribution of association p-values. Genes with at least one SNP association at p < 1 × 10⁻⁶ in the permuted analysis were flagged as potentially inflated and excluded from all downstream analyses.

The number of PEER factors was selected empirically for each cell type. For each candidate PEER factor count, ranging from 0 to 30 in increments of 5, we performed eQTL mapping on a representative subset of genes from chromosomes 1 and 19. We then jointly evaluated the number of permutation-flagged genes and the number of significant eQTL associations observed in the real data. The final PEER factor count for each cell type was selected to minimize inflation in the permuted data while preserving eGene discovery in the observed data, balancing model calibration with statistical power.

### Identification of primary and conditional single cell eQTL signals

eGenes were identified using gene-level p-values generated by the aggregated Cauchy association test (ACAT) from SAIGE-QTL step 3^26^. Genome-wide multiple testing correction was performed using Storey’s q-value method. Genes with q < 0.05 were considered significant eGenes. For each significant eGene, the variant with the smallest association p-value within the ±500 kb *cis*-window was designated as the primary sc-eQTL signal.

To identify additional independent single cell eQTL signals beyond the primary signal, we performed iterative conditional sc-eQTL analysis using SAIGE-QTL. For each significant eGene, the lead variant from the previous round was included as a covariate in subsequent association testing across the same *cis*-window. At each iteration, the variant with the smallest conditional association p-value was selected as a candidate secondary sc-eQTL signal if it passed a conditional significance threshold of p < 1 × 10⁻⁵. Iterative conditioning was repeated until no additional variants passed this threshold.

To further evaluate whether candidate conditional signals represented independent regulatory associations, we performed an all-but-one conditional analysis for genes with multiple detected single cell eQTL signals. For each target variant, association testing was repeated while conditioning on all other lead variants identified for the same gene, excluding the target variant itself. Variants that remained significant in this analysis at p < 1 × 10⁻⁵ were retained as independent conditional single cell eQTL signals.

### Gene constraint analysis

We downloaded gnomAD v4.1 gene constraint metrics from the gnomAD website ( https://gnomad.broadinstitute.org/) and extracted LOEUF and pLI scores to evaluate whether genes harboring conditional sc-eQTL signals differed in selective constraint^27,28^. Analyses were restricted to canonical transcripts and to genes with available constraint annotations. For each cell type, validated sc-eQTL signals were collapsed to the gene level and grouped according to the number of independent signals identified per gene. LOEUF distributions were compared across genes with one, two, or three or more independent sc-eQTL signals. As a complementary analysis, genes were grouped as having either one or two or more independent signals, and the proportion of highly constrained genes, defined as pLI ≥ 0.9, was calculated for each group.

### MashR and pairwise correlation analysis

To evaluate the extent of cell-type sharing among single cell eQTL effects, we applied mashr v0.2.73 separately to primary and conditional single cell eQTL signals^30^. For each pairwise cell-type comparison, we constructed a union set of significant SNP–gene pairs detected in both cell types and extracted the corresponding effect-size estimates and standard errors from the SAIGE-QTL summary statistics.

Pairwise mashr models were fit across all cell-type pairs to estimate sharing of effect direction and effect magnitude. Effects were considered shared in sign when the local false-sign rate was below the specified threshold using factor = 0, and shared in magnitude when effect estimates were consistent within a two-fold range using factor = 0.5.

### Fine-mapping of conditionally independent single cell eQTL signals

Independent single cell eQTL signals identified through iterative conditional analysis were fine-mapped using SuSiE-RSS v0.14.2^29^. In-sample LD matrices were calculated from the same genotype cohort used for eQTL mapping.

For each gene, we used round-specific SAIGE-QTL summary statistics for fine-mapping. Round 1 summary statistics were used to fine-map the primary sc-eQTL signal, whereas conditional summary statistics from subsequent rounds were used to fine-map secondary and higher-order signals. To isolate a single independent association signal at each iteration, SuSiE-RSS was run with L = 1 for each round-specific analysis. PIPs and CSs were estimated using variants within the corresponding *cis*-window.

Variants included in the resulting credible sets were considered candidate causal variants for each independent single cell eQTL signal.

### Colocalization Analysis

We performed colocalization analyses between independent sc-eQTL signals and GWAS summary statistics for AD^34^ or SCZ^35^ using coloc v6.0.1^36^.

For each gene, colocalization was performed separately for each independent sc-eQTL signal identified through iterative conditional analysis. Primary sc-eQTL signals were analyzed using the corresponding marginal eQTL summary statistics, whereas secondary and higher-order sc-eQTL signals were analyzed using conditional eQTL summary statistics from the relevant iteration. GWAS summary statistics were restricted to the same *cis*-window used for eQTL fine-mapping. Variants were matched across eQTL and GWAS datasets using chromosome, position, reference allele, and alternate allele information. Colocalization posterior probabilities for hypotheses H0–H4 were estimated using effect sizes, standard errors, sample sizes, and allele frequencies from both the eQTL and GWAS datasets.

### CUT&Tag peak calling and differential peak analysis

Raw CUT&Tag sequencing reads were obtained from Ziegler et al. and aligned to the human reference genome GRCh38 using bwa-mem with default parameters^31^. Peak calling was performed from aligned BAM files using MACS2 v2.2.7.1^45^, and resulting peak files were exported in BED format for downstream analyses and genome browser visualization.

For H3K27ac CUT&Tag datasets, peaks were called using parameters optimized for narrow regulatory elements: macs2 callpeak -q 0.00001 --narrow --nolambda --nomodel --shift -75 --extsize 150 --keep-dup all --format BAMPE. For H3K4me3 CUT&Tag datasets, peaks were called using broad-peak parameters: macs2 callpeak -q 0.01 --broad --broad-cutoff 0.05 --keep-dup all --format BAMPE.

Differential peak analyses were performed separately for H3K27ac and H3K4me3 using donor-aware count-based models implemented in DiffBind v3.10.1^46^. The dataset included six FANS-isolated cell populations: ERG+, NeuN+, NOTCH3+, OLIG2+, PU.1+, and RFX4+, corresponding to endothelial cells, neurons, mural cells, oligodendrocytes, microglia, and astrocytes, respectively. Each cell population included six biological donors.

For each histone modification, samples were grouped by antibody and cell type. Peaks overlapping regions in the ENCODE hg38 unified blacklist were removed before read counting^47^. Consensus peak sets were generated using dba.count() with a minimum overlap of three samples, and peak summits were recentered to 500-bp windows.

### Identification of cell type-specific and cell type-shared peaks

Pairwise differential binding analyses were performed across all cell type combinations using DESeq2 v1.40.2 as implemented in DiffBind^46,48^. For each histone modification, a generalized linear model including donor and cell type was fit for all contrasts. Peaks were considered significantly differential if they met an FDR < 0.05 after Benjamini-Hochberg correction and an absolute log_2_ fold change ≥ 1.

Cell type-specific peaks were defined as peaks significantly enriched in one cell type relative to each of the other five cell populations across all pairwise comparisons. Peaks satisfying these criteria for a given cell type were designated cell type-specific.

Cell type-shared peaks were identified by assessing consensus peak occupancy across cell types using BEDTools intersect v2.30.0^49^. Peaks present in at least three cell types and not classified as cell type-specific were designated shared peaks.

### Promoter and enhancer peak annotation

Peak sets were annotated using ChIPseeker v1.36.0 with TxDb.Hsapiens.UCSC.hg38.knownGene^50^. Promoter peaks were defined as H3K4me3 peaks located within 3 kb of any TSS. Putative enhancers were defined as H3K27ac peaks located more than 3 kb from any TSS and lacking an overlapping H3K4me3 signal. Annotated peak sets were exported in BED format for downstream analyses and genome browser visualization.

### Permutation-based analysis of single cell eQTL overlap with regulatory elements

To test whether fine-mapped sc-eQTLs were preferentially located within regulatory elements, we performed a GoShifter-inspired permutation enrichment analysis^32^. Analyses were performed for each eQTL cell type against CUT&Tag-derived regulatory peak sets from five brain cell types: astrocytes, endothelial cells, microglia, neurons, and oligodendrocytes, as described above. Excitatory and inhibitory neuron eQTLs were compared with neuron CUT&Tag peaks, and OPC and oligodendrocyte eQTLs were compared with oligodendrocyte CUT&Tag peaks.

Fine-mapped variants with PIP > 0.01 were retained for analysis. We further restricted the analysis to genes with both primary and conditional sc-eQTL signals. For each gene, we defined a 1-Mb *cis*-window centered on the transcription start site, corresponding to ±500 kb from the TSS, and extracted peaks from the relevant cell-type peak set that overlapped this window.

The observed overlap statistic was defined as the sum of PIPs for credible-set variants that overlapped any peak within the *cis*-window. To generate a null distribution, we performed 1,000 circular permutations. In each permutation, all peaks within the *cis*-window were shifted by a random offset while preserving their size and spacing. Peak coordinates were wrapped around the *cis-*window boundary, and peaks crossing the boundary were split into two fragments.

Empirical p-values were calculated as the proportion of permutations with an overlap statistic greater than or equal to the observed value. We also calculated z-scores as the difference between the observed overlap statistic and the mean permuted overlap statistic, divided by the standard deviation of the permuted values. Genomic overlaps were performed using GenomicRanges v1.62.1^51^.

## Data Availability

All data produced in the present study are available upon reasonable request to the authors

## Acknowledgements

This work is supported by an NIH/NIA K08 (K08-AG-086591), a Pilot Project award from the Penn Alzheimer’s Disease Research Center (1P30AG072979), and an Alzheimer’s Association Clinician Scientist Fellowship.

