## Supplementary Figures for "A large-scale single cell map of primary and conditional regulatory variation in the human brain"

**Supplementary Figure S1: Number of conditional single cell eQTLs by round prior to all-but-one validation.** a) Number of conditional sc-eQTLs for each cell type by round prior to all-but-one validation. b) Number of conditional sc-eQTLs for each cell type by round prior to all-but-one validation. Note that for panel b, y-axis is on $\log_{10}$ scale.

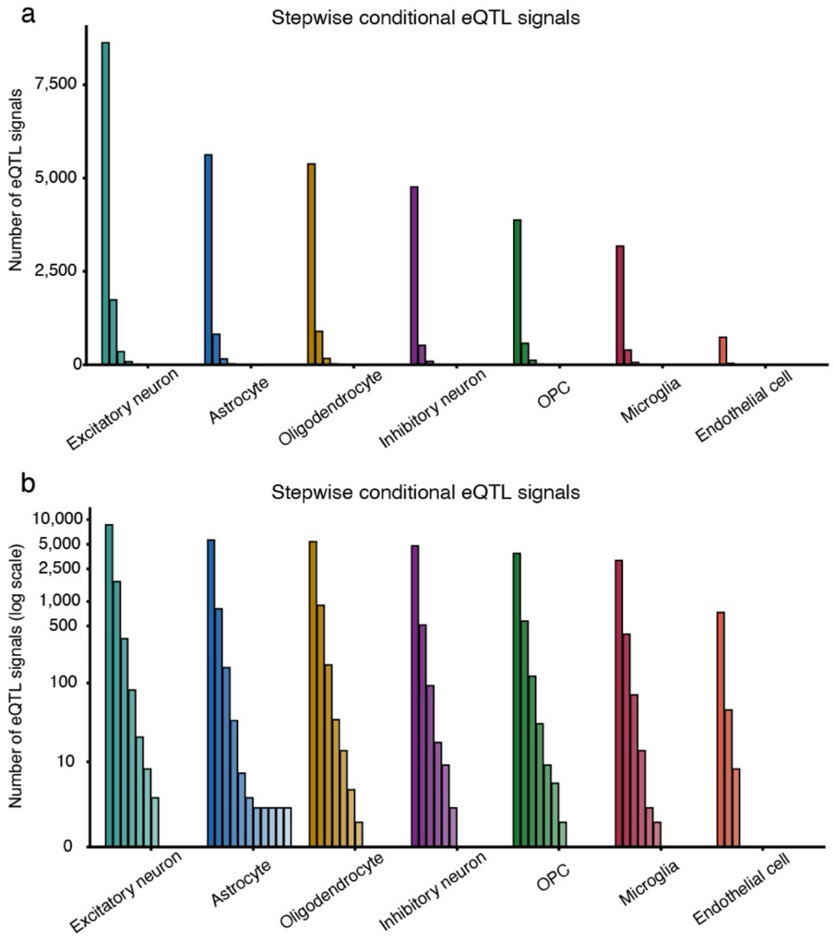

**Supplementary Figure S2: Number of conditional single cell eQTLs by round after all-but-one validation.**

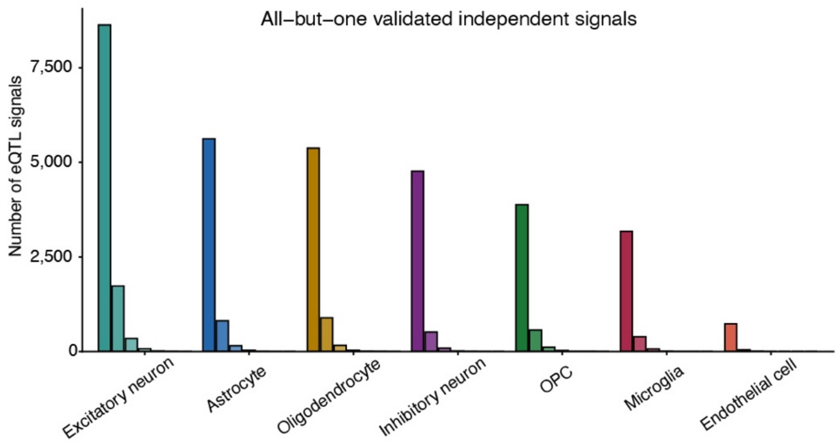

**Supplementary Figure S3: Proportion of eGenes with pL≥0.9 by cell type.**

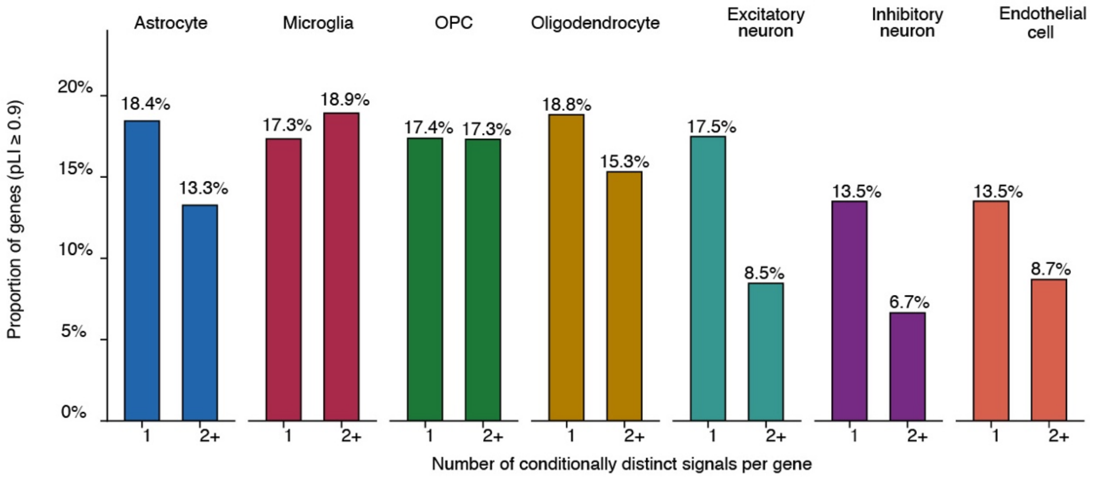

**Supplementary Figure 4: Cell type sharing of validated independent sc-eQTLs after all-but-one validation.** Upset plot shows the number of validated independent sc-eQTLs. Most sc-eQTLs were detected in a single cell type. eQTLs identified in multiple cell types are connected by lines.

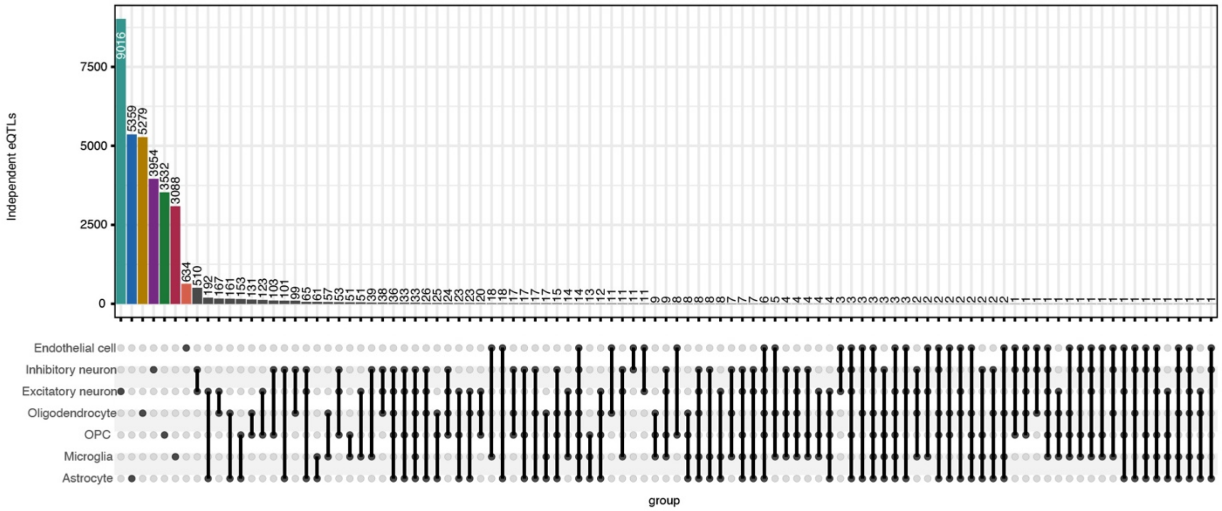

**Supplementary Figure 5: Mapping of cell type-specific sc-eQTLs.** a) Number of eQTL shared among cell types. Bar plot shows the number of eQTLs identified in a single cell type versus those shared between two or more cell types. b) Number of eGenes shared among cell types. Bar plot shows the number of eGenes identified in a single cell type versus those shared between two or more cell types. c) Expression of eGenes in multiple cell types. Bar plot shows the number of eGenes identified in a single cell type with at least one eQTL and their expression in one or more cell types. d) Sankey plot for the relationship between the cell types where eGenes are identified and the cell types where they are expressed.

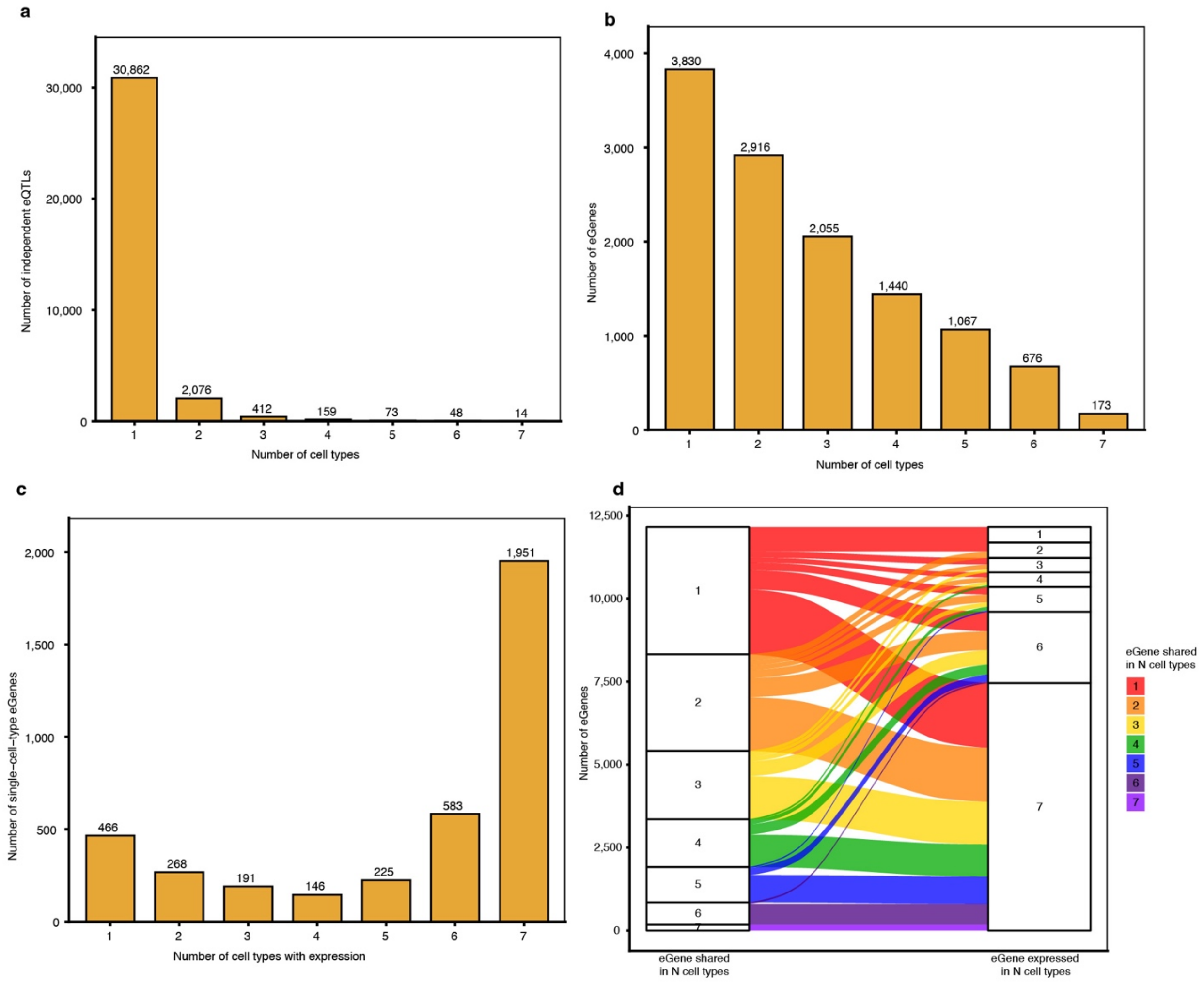

**Supplementary Figure S6: Regulation of multiple genes per single cell eQTL.** a) Number of eGenes per sc-eQTL signal, aggregated across all cell types. b) Number of eGenes per sc-eQTL, partitioned by cell type. c) Number of eGenes per primary sc-eQTL signal, aggregated across all cell types. d) Number of eGenes per conditional sc-eQTL signal, aggregated across all cell types.

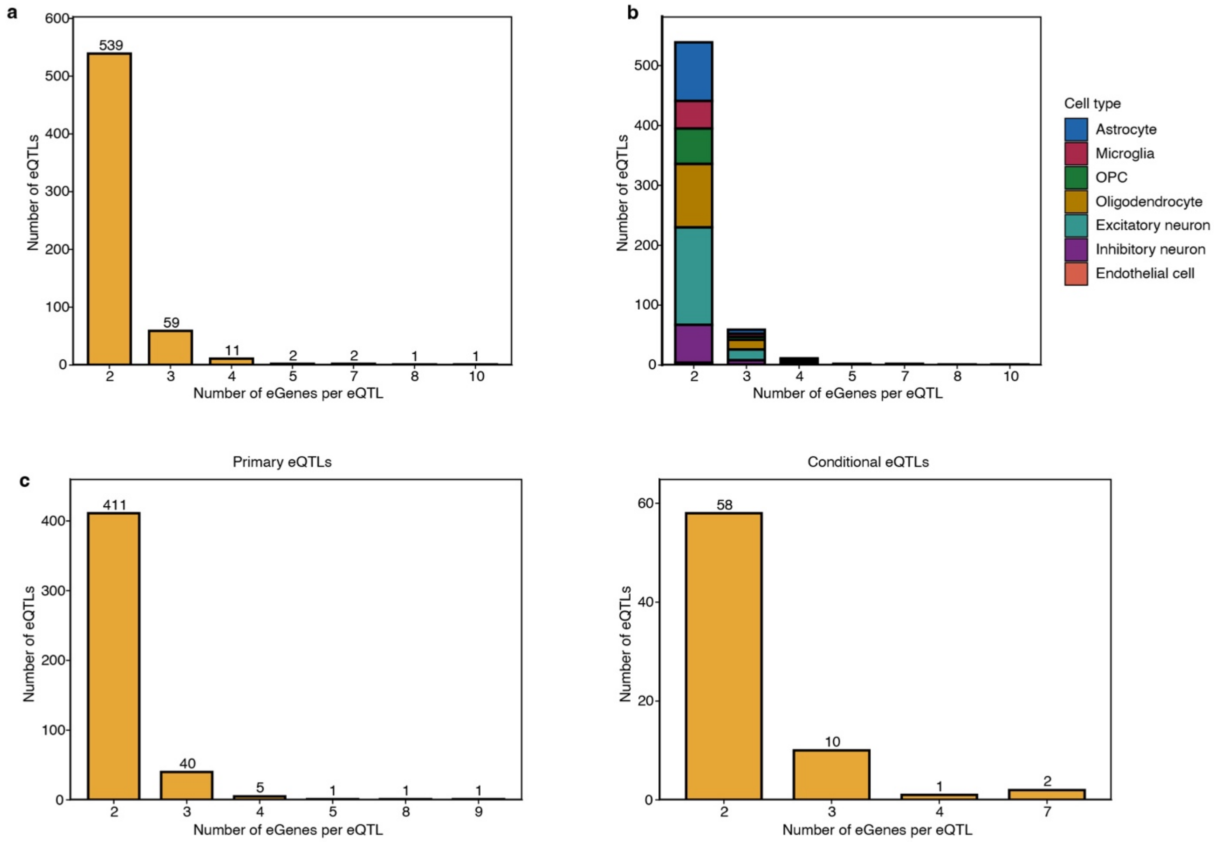

**Supplementary Figure S7: Pairwise sharing in effect direction for primary and conditional sc-eQTL signals.** Heatmaps show pairwise sharing in effect direction for primary and conditional sc-eQTL signals estimated using mashr (Urbut 2019). For each cell type pair, significant SNP-gene pairs detected in both cell types were analyzed. Values represent the proportion of effects shared in sign.

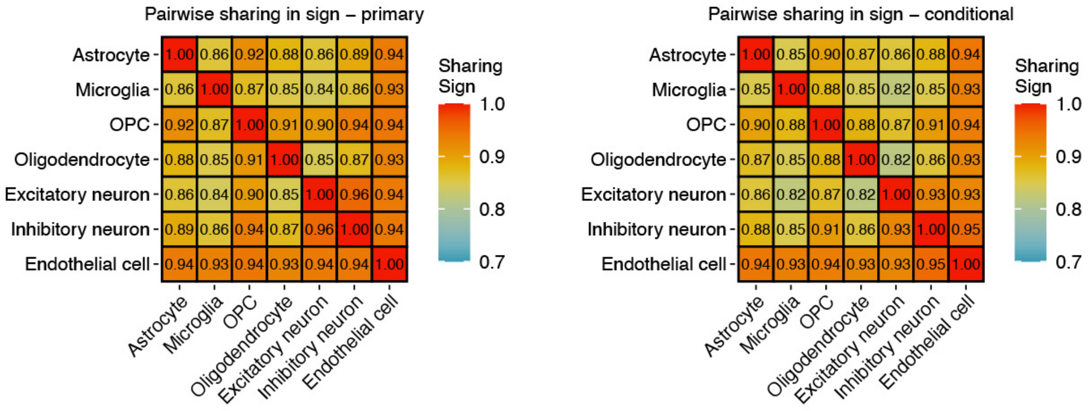

**Supplementary Figure S8: Distance to TSS for primary versus conditional single cell eQTLs.** Results are restricted to only eGenes with at least one conditional sc-eQTL in the cell type.

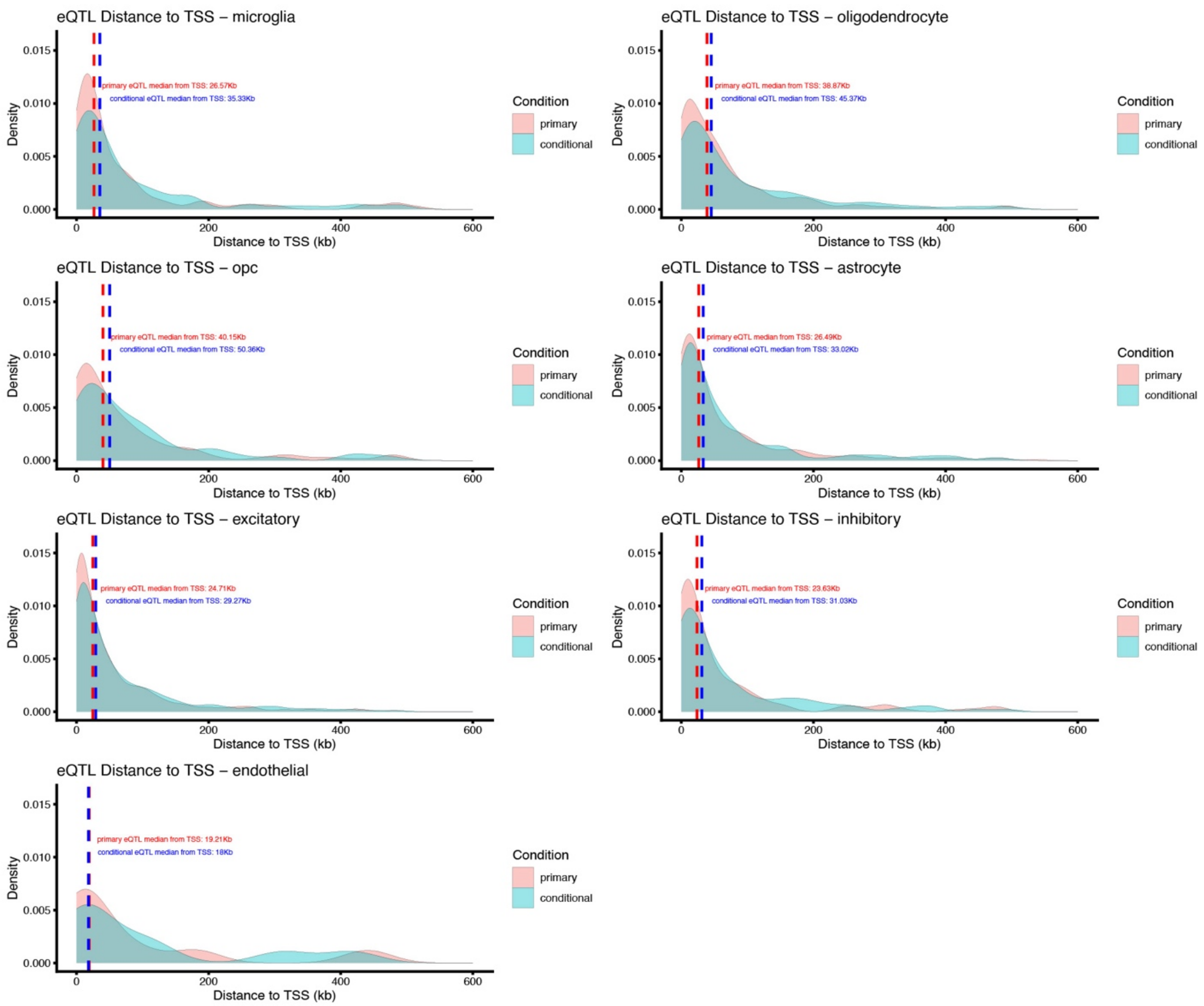

**Supplementary Figure S9: Distance to TSS for primary versus conditional single cell eQTLs by round.** sc-eQTLs are shown even if there are no conditional sc-eQTLs in that cell type.

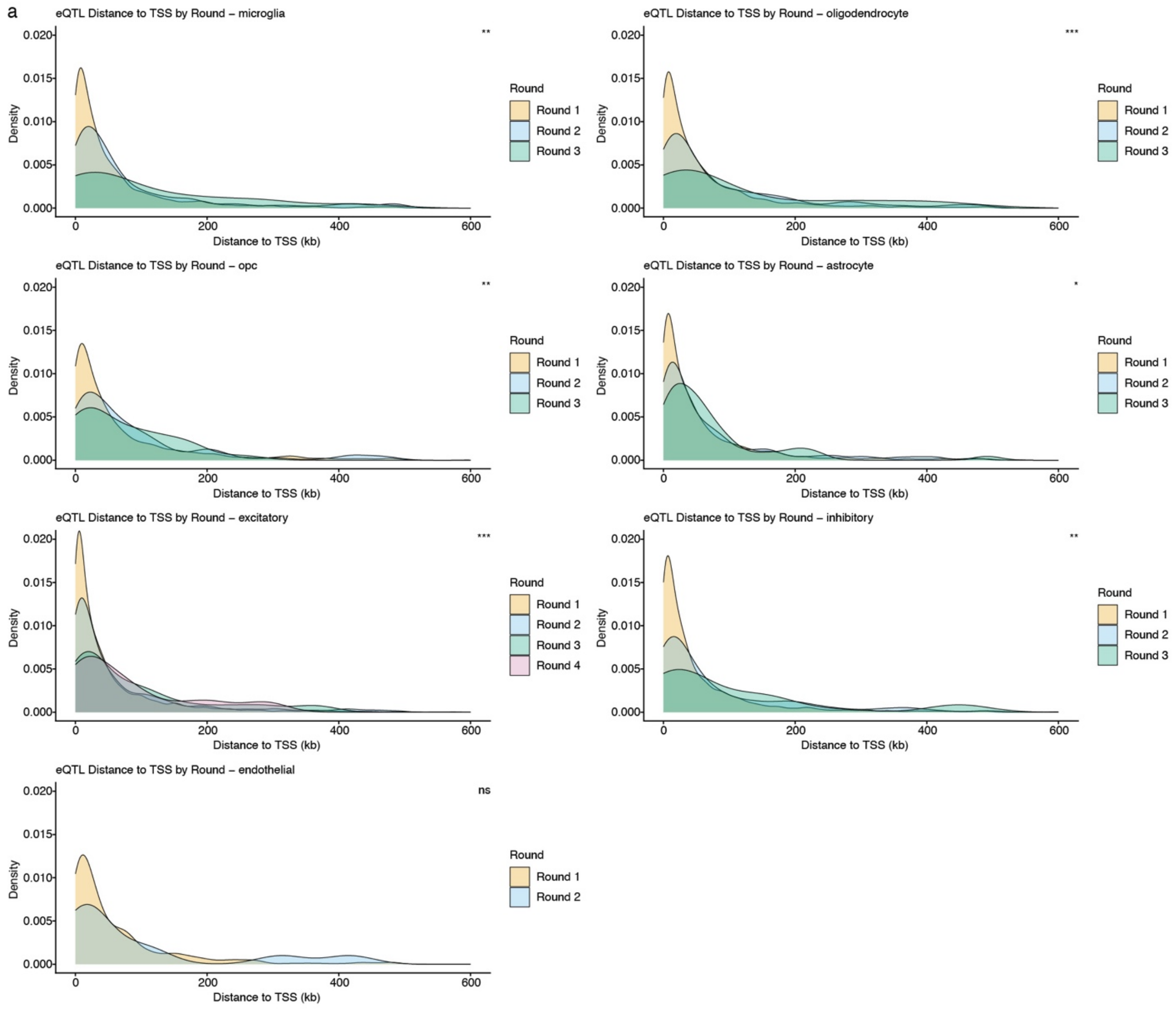

**Supplementary Figure S10: Genomic annotations primary versus conditional single cell eQTLs.** Results are restricted to only eGenes with at least one conditional sc-eQTL in the cell type.

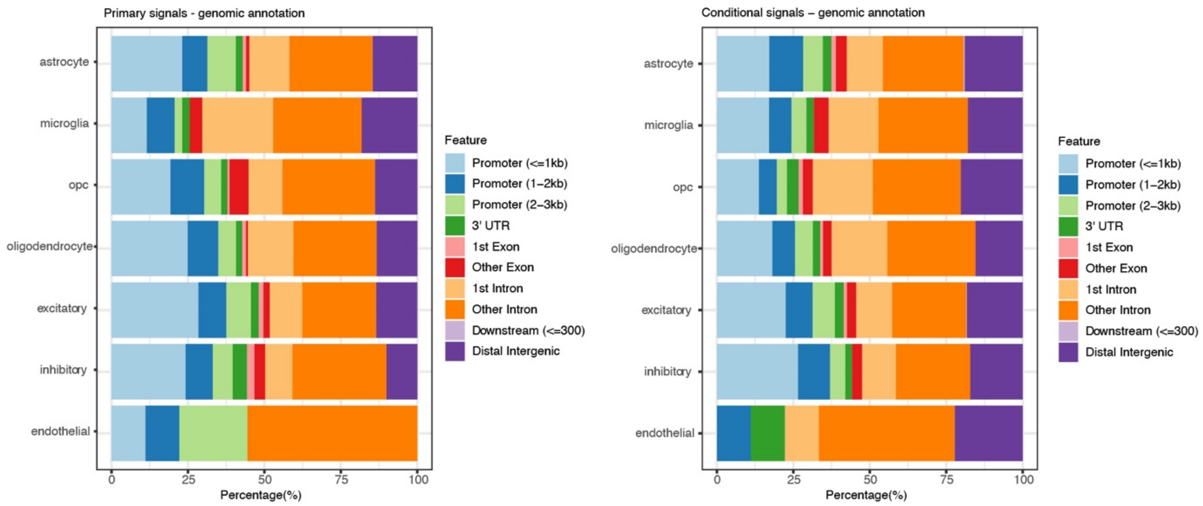

**Supplementary Figure S11: Number of colocalized genes at PP.H4>0.5 for AD and SCZ by cell type.**

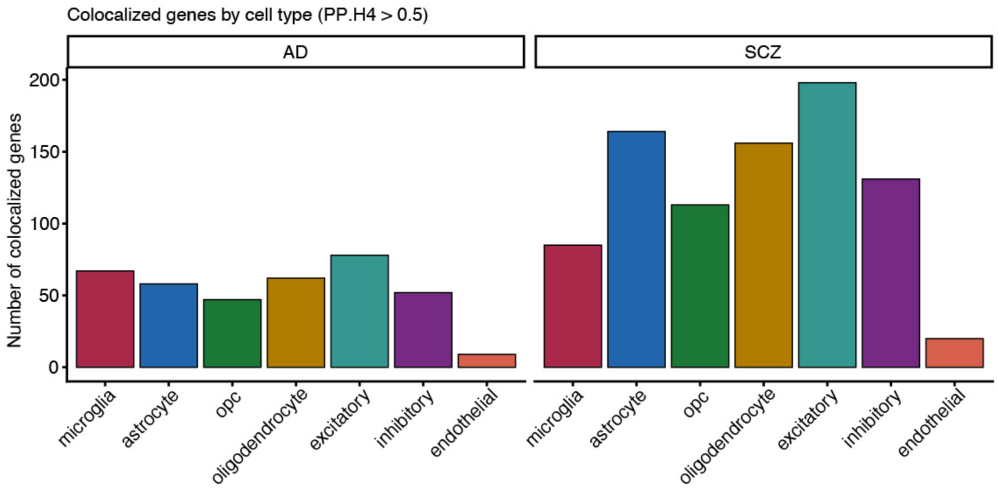
